# Reproductive Factors and Risk of Breast Cancer by Tumor Subtypes among Ghanaian Women: A Population-based Case-control Study

**DOI:** 10.1101/19006833

**Authors:** Jonine D Figueroa, Brittny C Davis Lynn, Lawrence Edusei, Nicholas Titiloye, Ernest Adjei, Joe-Nat Clegg-Lamptey, Joel Yarney, Beatrice Wiafe-Addai, Baffour Awuah, Maire A. Duggan, Seth Wiafe, Kofi Nyarko, Francis Aitpillah, Daniel Ansong, Stephen M Hewitt, Thomas Ahearn, Montserrat Garcia-Closas, Louise A Brinton, on behalf of the Ghana Breast Health Study team

**Affiliations:** Division of Cancer Epidemiology and Genetics, National Cancer Institute, Bethesda, MD, USA; Usher Institute and CRUK Edinburgh Centre, University of Edinburgh, Edinburgh, UK; Korle Bu Teaching Hospital, Accra, Ghana; Komfo Anokye Teaching Hospital, Kumasi, Ghana; Peace and Love Hospital, Kumasi, Ghana; Department of Pathology and Laboratory Medicine, University of Calgary, Calgary, Canada; Loma Linda University, School of Public Health, Loma Linda, CA, USA; University of Ghana, Accra, Ghana; Center for Cancer Research, National Cancer Institute, Bethesda, MD, USA

**Keywords:** Reproductive risk factors, subtype heterogeneity, racial disparities, breast cancer

## Abstract

**Background:** Higher proportions of early-onset and estrogen receptor (ER) negative cancers are observed in women of African ancestry than in women of European ancestry. Differences in risk factor distributions and associations by age at diagnosis and ER status may explain this disparity.

**Methods:** We analyzed data from 1,126 women (aged 18–74 years) with invasive breast cancer and 2,106 population controls recruited from three hospitals in Ghana from 2013 to 2015. Odds ratios (OR) and 95% confidence intervals (CI) were estimated for menstrual and reproductive factors using polytomous logistic regression models adjusted for potential confounders.

**Results:** Among controls, medians for age at menarche, parity, age at first birth, and breastfeeding/pregnancy were 15 years, 4 births, 20 years, and 18 months, respectively. For women ≥ 50 years, parity and extended breastfeeding were associated with decreased risks: >5 births vs. nulliparous, OR 0.40 (95% CI 0.20–0.83) and 0.71 (95% CI 0.51–0.98) for ≥19 vs. <13 breastfeeding months/pregnancy, which did not differ by ER. In contrast, for earlier onset cases (<50 years) parity was associated with increased risk for ER-negative tumors (*P*-heterogeneity by ER = 0.02), which was offset by extended breastfeeding. Similar associations were observed by intrinsic-like subtypes. Less consistent relationships were observed with ages at menarche and first birth.

**Conclusion:** Reproductive risk factor distributions are different from European populations but exhibited etiologic heterogeneity by age at diagnosis and ER status similar to other populations. Differences in reproductive patterns and subtype heterogeneity are consistent with racial disparities in subtype distributions.

**Key Messages:**

- Distribution of intrinsic-like breast cancer subtypes among Ghanaian women are distinct compared to European ancestry populations, with a higher proportion of ER-negative subtypes at younger ages.
- Increasing number of births and extended breastfeeding were associated with reduced risk for both ER-positive and ER-negative subtypes among later-onset breast cancer cases (women age ≥50 years).
- Extended breastfeeding offset a direct association that we observed of multiparity with early-onset (women age <50 years) ER-negative breast cancers.
- Number of births and breastfeeding duration are much higher in Ghanaian women compared to women of European ancestry and African Americans, however the relationships with risk are consistent when assessed by molecular subtype.

## Introduction

Reproductive factors have been well documented as key breast cancer risk factors with direct associations observed with early ages at menarche, nulliparity, late ages at first birth and limited breastfeeding. Breast cancer is a heterogeneous disease, with differential etiologic associations for tumor subtypes defined by estrogen receptor (ER), progesterone receptor (PR) and human epidermal growth factor receptor 2 (HER2) status.(1) Most of these results derive from studies on European ancestry populations. Similar investigations among African ancestry populations are crucial given the differences in demographic and risk factor distributions and their disproportionately high incidence of early-onset breast cancer and ER-negative aggressive subtypes.(1-4)

Analyses of risk factors by the African American Breast Cancer Epidemiology and Risk (AMBER) consortium have revealed differential risk factor associations by tumor subtypes defined by ER, PR, and HER2 status.(5,6) Parity was associated with a decreased risk for ER-positive cancers but an increased risk for triple-negative breast tumors; furthermore, ever breastfeeding in parous women was strongly inversely related to the risk of triple-negative tumors.(6) Accumulating data support similar observations in other studies on women of African American and European ancestry, although distributions of risk factors differ.(1,7-11)

With substantially increasing rates of breast cancer in sub-Saharan Africa, identifying risk factors and strategies for reducing incidence are essential.(12,13) A population-based case-control study of breast cancer in Ghana aimed to overcome challenges of previous African studies that were unable to select population-based controls and properly classify hormone receptor-negative cases.(3,12,14,15) Using a census-based sampling of controls (16) and standardized protocols for collecting tumor biopsy samples for immunohistochemical (IHC) staining from cases prior to treatment (17), we sought to determine the associations between menstrual and reproductive risk factors and breast cancer subtypes.

## Methods

See Supplemental Material for further details on methods for study population, risk factor information, ethical approvals, tumor characteristics, and statistical analysis.

### Study population

Details of the multi-disciplinary population-based case-control study in Ghana have been previously described.(17-18, 22) Our primary analyses focused on ER status because this was the key marker of etiological heterogeneity demonstrated in previous studies.(19)

### Statistical analysis

Odds ratios (ORs) and 95% confidence intervals (95% CIs) were estimated to determine menstrual and reproductive factors using polytomous logistic models adjusted for study site and age (as a categorical variable) as well as key risk factors, including education, a family history of breast cancer, body size (17) and menopausal status or age at menopause. All statistical tests were two-sided. Analyses were performed using STATA/MP 14.2 (StataCorp, College Station, TX). Plots on the means and standard deviations of a 3-point running average were obtained using R version 3.4.4.

## Results

### Descriptive characteristics of cases and controls

Cases were slightly older than controls reflecting that the controls were initially frequency matched to all women with a suspicion of breast cancer prior to diagnosis confirmation. Approximately half of these women had non-malignant breast diseases and tended to be younger than those with malignant breast disease.(17) The cases more often than the controls reported late ages at menarche, few births, late ages at first birth and low median breastfeeding months (Table 1).

**Table 1.**
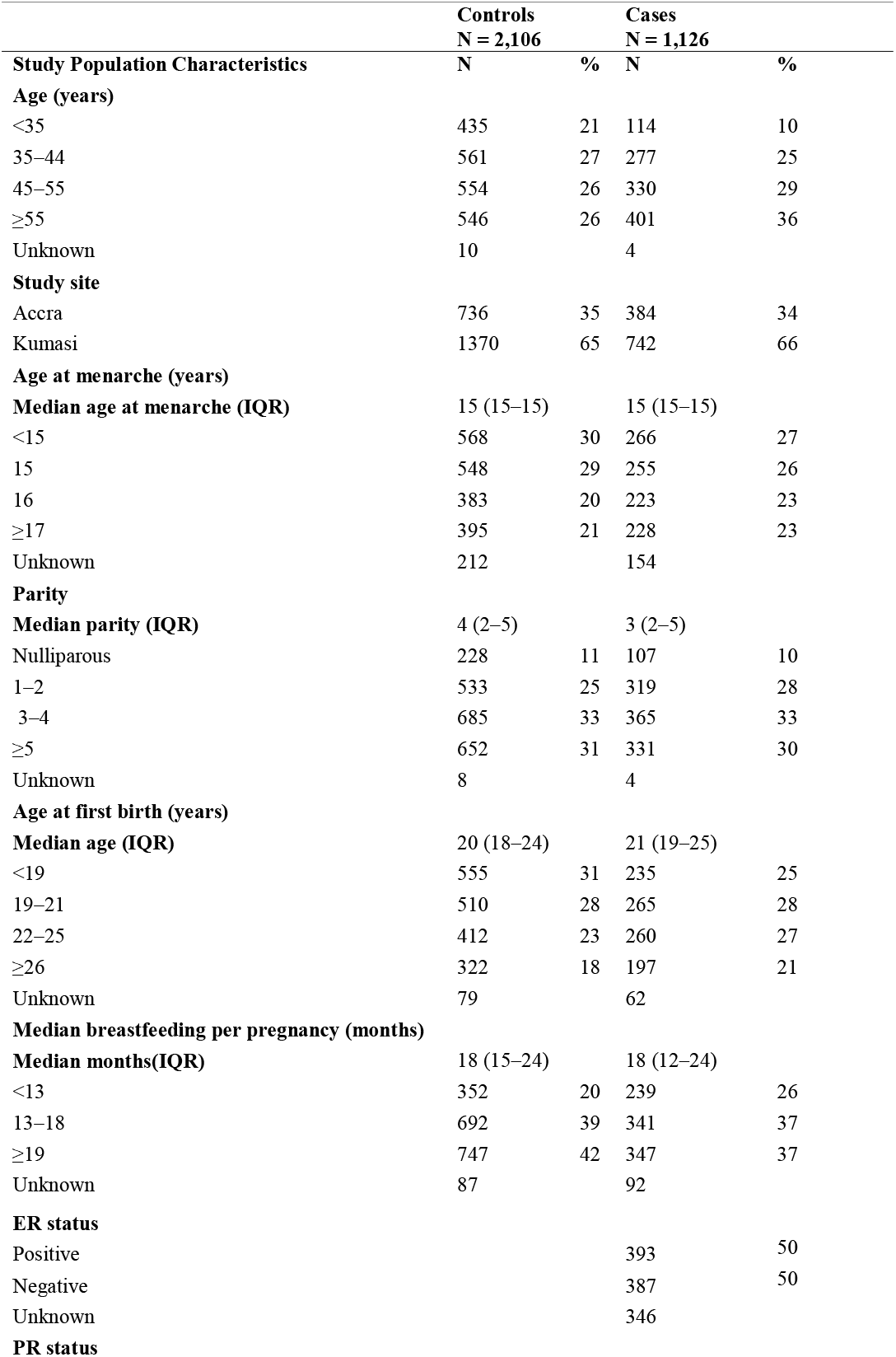
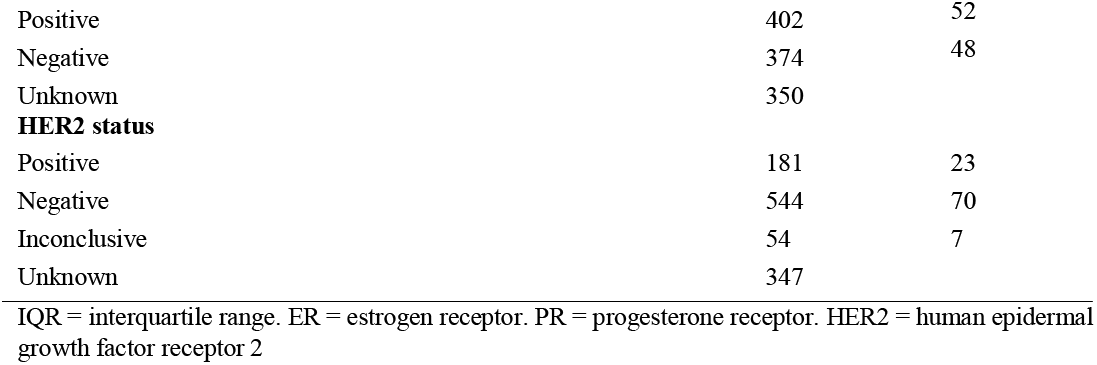
Demographic and reproductive characteristics of 1,126 diagnosed invasive breast cancer cases and 2,106 controls from the Ghana Breast Health Study.

| **Study Population Characteristics** | **Controls**<br>**N = 2,106** |  | **Cases**<br>**N = 1,126** |  |
| --- | --- | --- | --- | --- |
|  | **N** | **%** | **N** | **%** |
| **Age (years)** |  |  |  |  |
| <35 | 435 | 21 | 114 | 10 |
| 35–44 | 561 | 27 | 277 | 25 |
| 45–55 | 554 | 26 | 330 | 29 |
| ≥55 | 546 | 26 | 401 | 36 |
| Unknown | 10 |  | 4 |  |
| **Study site** |  |  |  |  |
| Accra | 736 | 35 | 384 | 34 |
| Kumasi | 1370 | 65 | 742 | 66 |
| **Age at menarche (years)** |  |  |  |  |
| **Median age at menarche (IQR)** | 15 (15–15) |  | 15 (15–15) |  |
| <15 | 568 | 30 | 266 | 27 |
| 15 | 548 | 29 | 255 | 26 |
| 16 | 383 | 20 | 223 | 23 |
| ≥17 | 395 | 21 | 228 | 23 |
| Unknown | 212 |  | 154 |  |
| **Parity** |  |  |  |  |
| **Median parity (IQR)** | 4 (2–5) |  | 3 (2–5) |  |
| Nulliparous | 228 | 11 | 107 | 10 |
| 1–2 | 533 | 25 | 319 | 28 |
| 3–4 | 685 | 33 | 365 | 33 |
| ≥5 | 652 | 31 | 331 | 30 |
| Unknown | 8 |  | 4 |  |
| **Age at first birth (years)** |  |  |  |  |
| **Median age (IQR)** | 20 (18–24) |  | 21 (19–25) |  |
| <19 | 555 | 31 | 235 | 25 |
| 19–21 | 510 | 28 | 265 | 28 |
| 22–25 | 412 | 23 | 260 | 27 |
| ≥26 | 322 | 18 | 197 | 21 |
| Unknown | 79 |  | 62 |  |
| **Median breastfeeding per pregnancy (months)** |  |  |  |  |
| **Median months(IQR)** | 18 (15–24) |  | 18 (12–24) |  |
| <13 | 352 | 20 | 239 | 26 |
| 13–18 | 692 | 39 | 341 | 37 |
| ≥19 | 747 | 42 | 347 | 37 |
| Unknown | 87 |  | 92 |  |
| **ER status** |  |  |  |  |
| Positive |  |  | 393 | 50 |
| Negative |  |  | 387 | 50 |
| Unknown |  |  | 346 |  |
| **PR status** |  |  |  |  |

|  |  |  |
| --- | --- | --- |
| Positive | 402 | 52 |
| Negative | 374 | 48 |
| Unknown | 350 |  |
| **HER2 status** |  |  |
| Positive | 181 | 23 |
| Negative | 544 | 70 |
| Inconclusive | 54 | 7 |
| Unknown | 347 |  |
| IQR = interquartile range. ER = estrogen receptor. PR = progesterone receptor. HER2 = human epidermal growth factor receptor 2 |  |  |

A total of 50%, 52% and 23% of cases were ER positive, PR positive, and HER2 positive, respectively (Table 1). Luminal A-like breast cancer was the most common subtype (49%) followed by triple-negative/basal-like (28%), HER2-enriched (15%) and luminal B-like breast cancers (8%) (Fig. 1). There were no significant differences in cases missing ER, PR and HER2 status by risk factor data (data not shown).

**Fig. 1:**
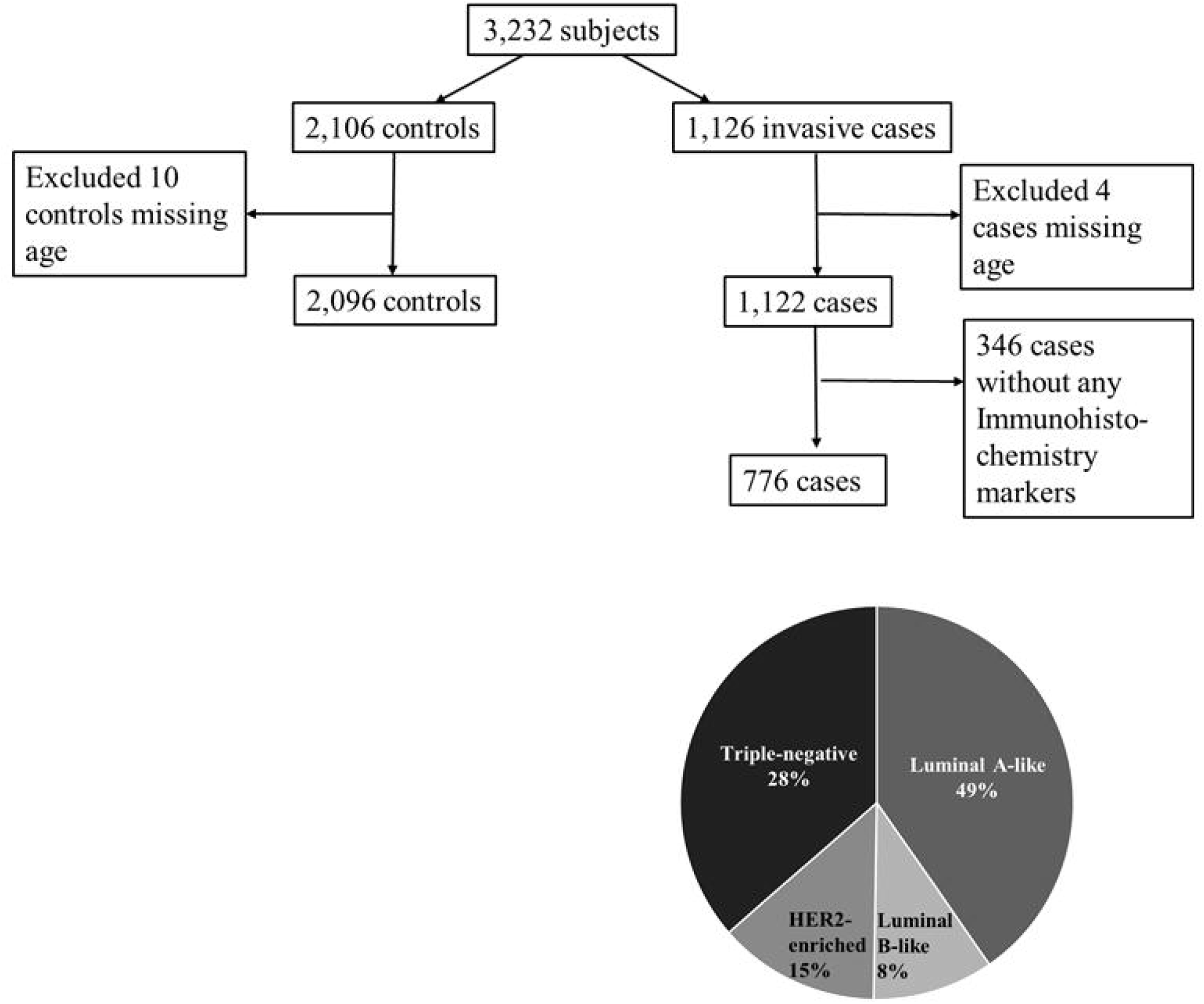
Details of cases and controls for analysis of reproductive factors and breast cancer risk by tumor characteristics in the Ghana Breast Health Study. Intrinsic-like subtype distribution for cases with immunohistochemical data on ER, PR, and HER2 defined as luminal A-like (ER+ or PR+ and HER2−), luminal B-like (ER+ or PR+ and HER2+), HER2-enriched-like (ER− or PR− and HER2+), or triple-negative/basal-like (ER−, PR− and HER2−).

We assessed descriptively if reproductive factors varied by birth cohort (1945–1975), which are presented in Fig. 2. Number of births was lower in later birth cohorts compared to earlier birth cohorts, with cases having fewer births on average compared with the controls (1945, mean 4.1 for cases and 6.4 for controls; 1975, mean 2.5 for cases and 3.3 for controls). Age at first birth increased by approximately 1 year in the later compared with earlier birth cohorts for both cases and controls (21.7 in 1945 and 22.3 in 1975 among controls). Age at menarche showed no apparent trends, hovering around 15 years across the birth cohorts. Breastfeeding months per pregnancy among controls declined until the 1960s and steadily increased until 1975. Among the cases, breastfeeding months per pregnancy increased over time by 1 month per pregnancy from 17 to 18 months.

**Fig. 2:**
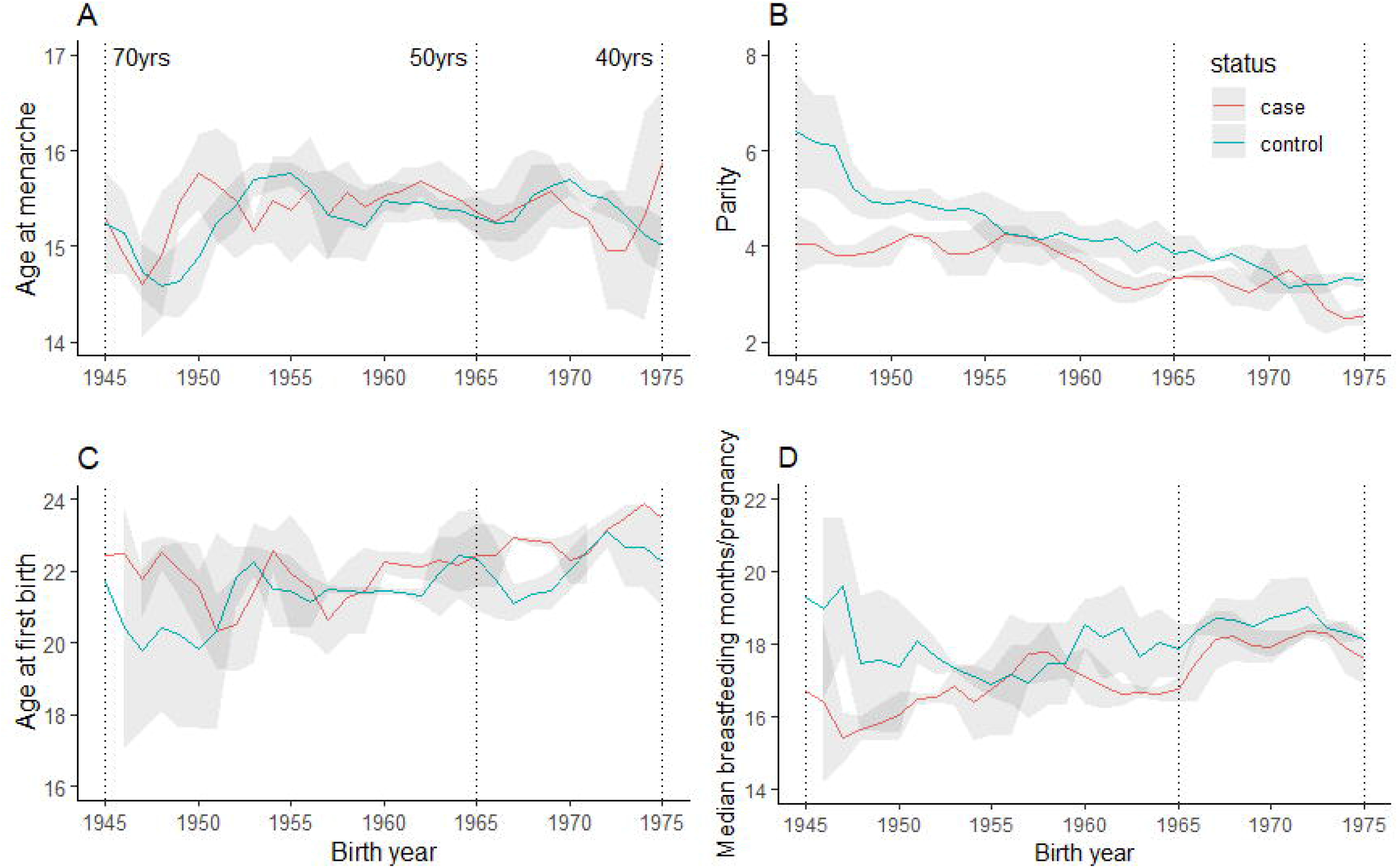
Temporal trends of menstrual and reproductive risk factors for cases and controls in the Ghana Breast Health Study by birth cohorts from 1945 to 1975. (A) Age at menarche, (B) parity, (C) age at first birth and (D) median breastfeeding months per pregnancy. The means and standard deviations plotted are the results of a 3-point running average. Gray indicates standard deviation.

### Associations with reproductive factors overall and stratified by age

Associations for age at menarche, number of births, age at first birth, and median breastfeeding months per pregnancy overall and stratified by age are shown in Table 2. Analyses of all cases combined showed number of births as the only risk factor with a statistically significant risk association (*P* -trend=0.005). Among women aged <50 years, we observed an inverse association with parity (≥5 vs 0 births: OR 0.70, 95% CI 0.42–1.18, *P* - trend = 0.06) and an increased risk with older ages at first birth (≥26 vs <19 years: OR 1.40, 95% CI 0.97–2.01, *P* -trend = 0.05). In more discrete categories of age, we observed a significant trend (*P* = 0.01) with advancing age at first birth among women aged <40 years (Supplementary Table 1). Age at menarche and median breastfeeding months were not significantly associated with breast cancer risk among younger women. Among women aged ≥50 years, a strong inverse association was observed with parity (≥5 vs 0 births: OR 0.40, 95% CI 0.20–0.83); a test for interaction with age was significant (*P* = 0.02). Similarly, median breastfeeding months among older women were inversely associated with risk (≥19 vs <13 months: OR 0.71, 95% CI 0.51–0.98) and demonstrated a significant interaction with age (*P* = 0.01). Age at menarche was unrelated to risk among the older women (Table 2). Evaluation of these associations with more detailed categories of age revealed a significant interaction by age for parity and median breastfeeding months per pregnancy, with the strongest inverse associations of parity and extended breastfeeding among women aged ≥60 years (Supplementary Table 1).

**Table 2.**
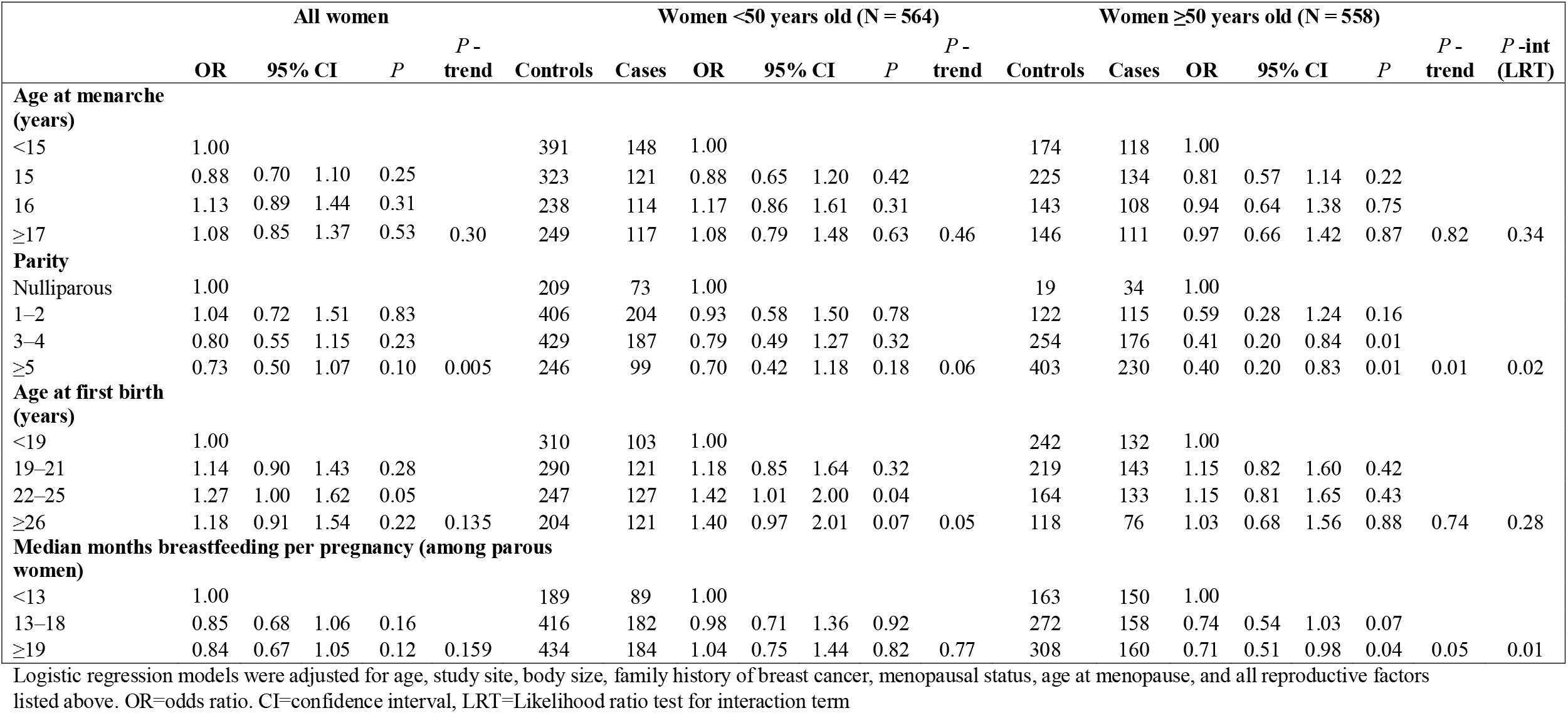
ORs and 95% CIs for select reproductive risk factors and breast cancer risk overall, stratified by women younger and older than 50 years among 1,122 cases and 2,096 controls.

### Associations with reproductive factors by ER and stratified by age

Analyses for all cases combined did not show statistically significant differences for ER-negative compared with ER-positive cases (Supplementary Table 2). When we evaluated the associations by ER status among women aged <50 years (Table 3), we observed a strong inverse association with parity for ER-positive tumors and a positive association for ER-negative tumors, with the test for heterogeneity being statistically significant (*P*-het = 0.02). Among women <50 years, older ages at first birth showed a slightly stronger association with increased risk for ER-positive than ER-negative breast tumors, but the test for heterogeneity was not statistically significant. Extended breastfeeding only showed an inverse association among ER-negative tumors, with evidence of significant heterogeneity compared with ER-positive tumors (≥19 vs <13 months: ER-positive tumors OR 1.39, 95% CI 0.82–2.34; ER-negative tumors 0.71, 0.45–1.12; *P*-het = 0.04). There was no additional relationship for ≥19 breastfeeding months; when we compared women with ≥13 breastfeeding months per pregnancy to <13 months, the resultant OR for ER-negative tumors was OR 0.69 (95% CI 0.45–1.03). There was a suggestion of a positive association with older ages at menarche for ER-positive breast tumors that was not apparent for ER-negative breast tumors.

**Table 3.** Reproductive risk factors and breast cancer risk stratified by ER status among women <50 years of age among 378 cases and 1,294 controls.

|  | ER-positive<br>N = 185 |  |  |  |  |  | ER-negative<br>N = 193 |  |  |  | ER-negative/ER-positive |  |
| --- | --- | --- | --- | --- | --- | --- | --- | --- | --- | --- | --- | --- |
|  | Controls | ER-positive | ER-negative | OR | 95% CI |  | P -<br>trend | OR | 95% CI |  | P -<br>trend | P-het |
| Age at menarche (years) |  |  |  |  |  |  |  |  |  |  |  |  |
| <15 | 391 | 40 | 54 | 1.00 |  |  |  | 1.00 |  |  |  |  |
| 15 | 323 | 44 | 37 | 1.21 | 0.76 | 1.93 |  | 0.69 | 0.43 | 1.09 |  | 0.08 |
| 16 | 238 | 37 | 43 | 1.38 | 0.85 | 2.25 |  | 1.21 | 0.77 | 1.90 |  | 0.79 |
| ≥17 | 249 | 47 | 38 | 1.61 | 1.00 | 2.58 | 0.05 | 0.97 | 0.61 | 1.55 | 0.81 | 0.09 |
| Parity |  |  |  |  |  |  |  |  |  |  |  |  |
| Nulliparous | 209 | 23 | 19 | 1.00 |  |  |  | 1.00 |  |  |  |  |
| 1–2 | 406 | 62 | 64 | 0.57 | 0.26 | 1.22 |  | 1.70 | 0.82 | 3.51 |  | 0.06 |
| 3–4 | 429 | 66 | 67 | 0.51 | 0.24 | 1.09 |  | 1.62 | 0.78 | 3.36 |  | 0.04 |
| ≥5 | 246 | 34 | 43 | 0.46 | 0.20 | 1.06 | 0.19 | 1.80 | 0.82 | 3.95 | 0.32 | 0.02 |
| Age at first birth (years) |  |  |  |  |  |  |  |  |  |  |  |  |
| <19 | 310 | 31 | 40 | 1.00 |  |  |  | 1.00 |  |  |  |  |
| 19–21 | 290 | 45 | 40 | 1.43 | 0.86 | 2.36 |  | 1.08 | 0.66 | 1.76 |  | 0.54 |
| 22–25 | 247 | 37 | 52 | 1.34 | 0.78 | 2.30 |  | 1.64 | 1.01 | 2.65 |  | 0.49 |
| ≥26 | 204 | 45 | 32 | 1.72 | 0.99 | 2.97 | 0.08 | 1.15 | 0.66 | 1.99 | 0.30 | 0.29 |
| Median months breastfeeding per pregnancy (among parous women) |  |  |  |  |  |  |  |  |  |  |  |  |
| <13 | 189 | 24 | 39 | 1.00 |  |  |  | 1.00 |  |  |  |  |
| 13–18 | 416 | 65 | 57 | 1.37 | 0.82 | 2.29 |  | 0.67 | 0.42 | 1.05 |  | 0.02 |
| ≥19 | 434 | 64 | 61 | 1.39 | 0.83 | 2.34 | 0.29 | 0.71 | 0.45 | 1.12 | 0.25 | 0.04 |

Polytomous logistic regression models were adjusted for age, education, study site, body size, family history of breast cancer, menopausal status, and all reproductive factors listed above. ER=estrogen receptor. OR=odds ratio. CI=confidence interval. P-het= *P*-heterogeneity test

Among the women aged ≥50 years (Table 4), parity was inversely associated with risk for both ER-negative and ER-positive tumors (although there were few nulliparous women, p-het = 0.33). Although extended breastfeeding showed an inverse association regardless of ER status, a stronger association was observed among ER-positive tumors (*P*-het = 0.07). Age at first birth did not demonstrate any consistent associations with risk.

**Table 4.** Risk factor associations stratified by ER status among women ≥50 years of age among 398 cases and 802 controls.

|  | ER-positive<br>N = 205 |  |  |  |  |  | ER-negative<br>N = 193 |  |  |  | ER-negative/ER-positive |  |
| --- | --- | --- | --- | --- | --- | --- | --- | --- | --- | --- | --- | --- |
|  | Controls | ER-positive | ER-negative | OR | 95% CI |  | P-trend | OR | 95% CI |  | P -<br>trend | P-het |
| Age at menarche (years) |  |  |  |  |  |  |  |  |  |  |  |  |
| <15 | 177 | 51 | 39 | 1.00 |  |  |  | 1.00 |  |  |  |  |
| 15 | 225 | 50 | 46 | 0.69 | 0.43 | 0.85 |  | 0.85 | 0.51 | 1.39 |  | 0.60 |
| 16 | 145 | 37 | 27 | 0.73 | 0.43 | 0.69 |  | 0.69 | 0.39 | 1.22 |  | 0.84 |
| ≥17 | 146 | 42 | 43 | 0.84 | 0.50 | 1.13 | 0.74 | 1.13 | 0.67 | 1.92 | 0.77 | 0.36 |
| Parity |  |  |  |  |  |  |  |  |  |  |  |  |
| Nulliparous | 19 | 12 | 14 | 1.00 |  |  |  | 1.00 |  |  |  |  |
| 1–2 | 127 | 48 | 36 | 0.82 | 0.31 | 2.16 |  | 0.34 | 0.13 | 0.88 |  | 0.13 |
| 3–4 | 256 | 72 | 53 | 0.58 | 0.23 | 1.49 |  | 0.23 | 0.09 | 0.57 |  | 0.83 |
| ≥5 | 406 | 76 | 91 | 0.49 | 0.19 | 1.26 | 0.24 | 0.28 | 0.11 | 0.70 | 0.004 | 0.33 |
| Age at first birth (years) |  |  |  |  |  |  |  |  |  |  |  |  |
| <19 | 245 | 46 | 50 | 1.00 |  |  |  | 1.00 |  |  |  |  |
| 19–21 | 220 | 49 | 56 | 1.10 | 0.67 | 1.78 |  | 1.26 | 0.80 | 2.00 |  | 0.55 |
| 22–25 | 165 | 48 | 42 | 1.10 | 0.66 | 1.83 |  | 1.06 | 0.64 | 1.75 |  | 1.00 |
| ≥26 | 118 | 32 | 24 | 1.09 | 0.61 | 1.93 | 0.72 | 1.02 | 0.57 | 1.84 | 0.95 | 0.84 |
| Median months breastfeeding per pregnancy (among parous women) |  |  |  |  |  |  |  |  |  |  |  |  |
| <13 | 163 | 68 | 42 | 1.00 |  |  |  | 1.00 |  |  |  |  |
| 13–18 | 276 | 54 | 53 | 0.61 | 0.39 | 0.95 |  | 0.81 | 0.50 | 1.30 |  | 0.35 |
| ≥19 | 313 | 51 | 66 | 0.54 | 0.34 | 0.85 | 0.01 | 0.89 | 0.56 | 1.42 | 0.82 | 0.07 |

Polytomous logistic regression models were adjusted for age, education, study site, body size, family history of breast cancer, menopausal status, age at menopause, and all reproductive factors listed above. ER=estrogen receptor. OR=odds ratio. CI=confidence index. *P*-het= *P*-heterogeneity test

We further assessed the joint effects of parity and breastfeeding per pregnancy (Fig. 3). Among women aged ≥50 years, increasing parity, and breastfeeding were associated with reduced risks for both ER-negative and ER-positive tumors, with the lowest risks observed among women with ≥3 births who breastfed for ≥13 months/pregnancy compared with nulliparous women (ER-negative cases: OR 0.45, 95% CI 0.21–0.95; ER-positive cases: OR 0.31, 95% CI 0.13–0.75). This trend was less apparent among women aged <50 years with ER-positive tumors [≥3 births who breastfed for ≥13 months/pregnancy compared with nulliparous women (OR 0.69, 95% CI 0.36–1.30)]. In contrast, among women aged <50 years with ER-negative tumors, compared with nulliparous women, the highest risk was for those with ≥3 births who breastfed <13 months/pregnancy (OR 1.91, 95% CI 0.89–4.10).Women with ≥3 births who breastfed, on average, ≥13 months per pregnancy were not at increased risk (OR 1.09, 95% CI 0.56–2.10), due to the multiplicative joint association of two factors associated with risk in opposite directions.

**Fig. 3:**
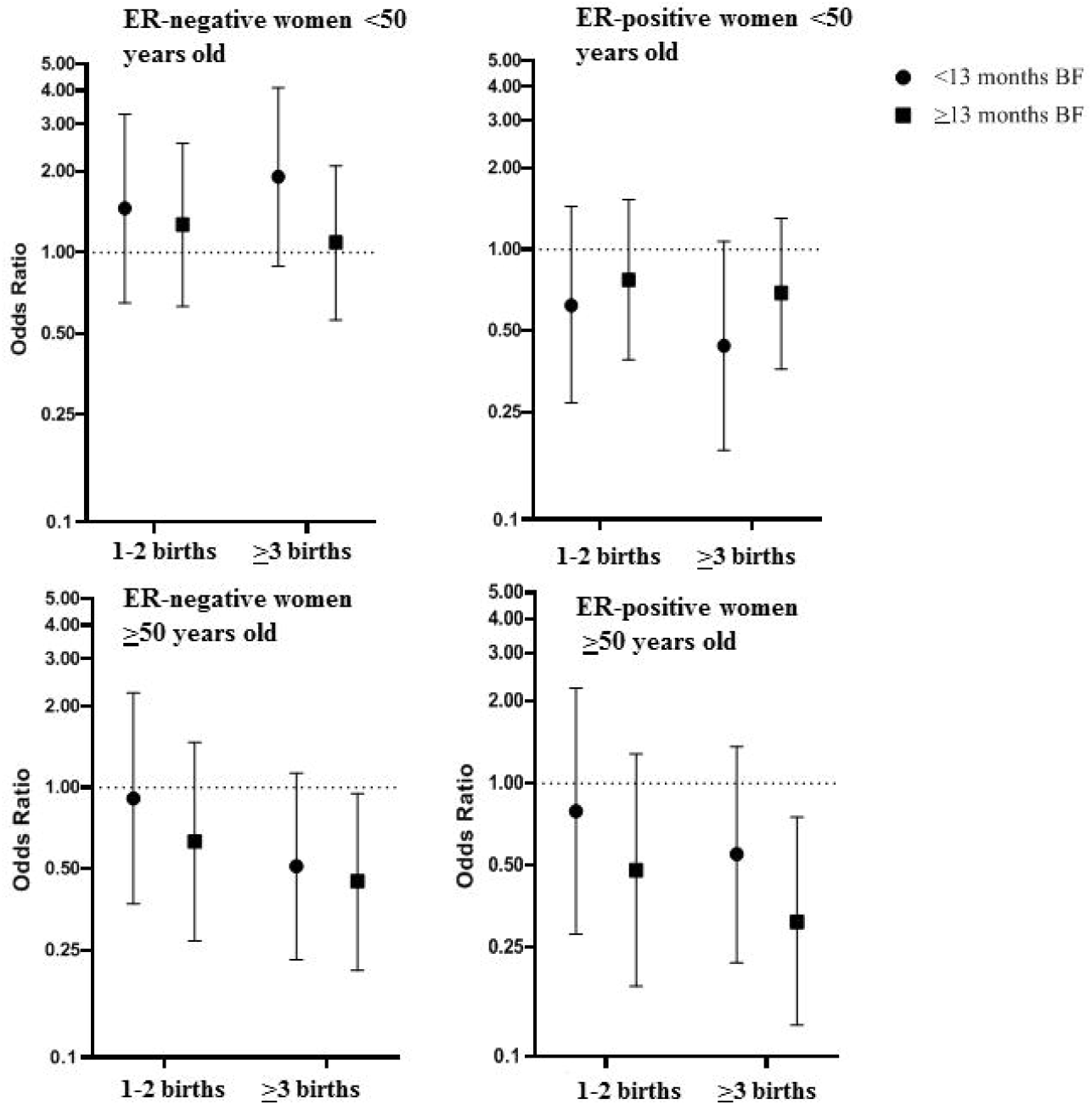
ORs and 95% CIs for joint effects of parity and breastfeeding (vs nulliparous) by ER status and age of onset. Polytomous logistic regression models were used to calculate ORs and 95% CI, adjusted for age, education, study site, body size, family history of breast cancer, age at menarche, age at first birth, menopausal status, and age at menopause. Error bars indicate standard deviations = breastfeeding. ER = Estrogen receptor.

### Associations with reproductive factors by ER, PR, HER2 status and stratified by age

We evaluated if associations with parity and breastfeeding differed using the IHC proxy for intrinsic subtypes. We focused our analyses on triple-negative compared with luminal A-like cases because previous studies have shown differences between these two groups (6-9,20,21) and these were also the two most common tumor subtypes (Supplementary Tables 3–4). Parity was inversely related to the risk of luminal A-like tumors regardless of age, as well as with risk of triple-negative tumors among women aged ≥50 years (Supplementary Tables 3–4). In contrast, a positive association was observed for triple-negative tumors among women aged <50 years (*P*-het = 0.03). Among younger women with triple-negative tumors, extended breastfeeding was inversely associated with risk, a relationship not observed for luminal A-like tumors. In contrast, among older women, we observed a strong inverse association of breastfeeding with luminal A-like tumors (OR 0.52, 95% CI 0.33–0.82) that was not observed for triple-negative tumors (*P* -het = 0.04) (Supplementary Table 4).

## Discussion

Among Ghanaian women, we observed substantial heterogeneity of the parity association with breast cancer risk by age at diagnosis and ER status, with strong inverse associations for all tumor subtypes in older (≥50 years) women and for younger-onset ER-positive tumors, but an opposite association for younger-onset ER-negative tumors (i.e. increased risk with increasing birth numbers). Median breastfeeding months per pregnancy was strongly inversely associated with later-onset breast tumor risk (particularly ER-positive or luminal A-like tumors); among younger women, it was an apparent protective factor for ER-negative tumors only. Similar to previous reports,(6-9,20,21) our study population allowed an evaluation of associations for a wide range of number of births and breastfeeding months per pregnancy.

Few studies have addressed the relation of reproductive risk factors in women of African ancestry. The largest dataset derives from the African Breast Cancer Study (ABCS),(22) a hospital-based case-control study in Nigeria, Cameroon and Uganda, comprising 1,995 cases and 2,631 controls (with 81% of the cases from Nigeria). Analyses from this study showed changing reproductive patterns over time (particularly number of births) and an inverse association of risk with parity; however, it did not show statistically significant heterogeneity of risk associations by menopausal status or age at diagnosis.(22,23) Notably, in contrast to our study, ABCS was not population-based and lacked information on hormone receptor status of the tumors, thereby limiting the comparability of the findings. Data from the AMBER consortium, a pooled analysis of four studies of African-American women with available tumor IHC data found that among 1,252 ER-negative breast tumors parous women were at elevated risk compared with nulliparous women, increasing to 1.60 among those aged <40 years.(6) Our data are consistent with AMBER and other recent studies,(9,11) supporting a cross-over association between parity on breast cancer risk that is dependent on age at onset and ER status.

In our Ghanaian population, number of births and breastfeeding years were highly correlated. Our data showed a significant inverse risk relationship with median breastfeeding months per pregnancy, with a 15% reduced risk for those with 13–18 vs <13 months/pregnancy. In pooled analyses of populations of European ancestry, breastfeeding has been shown to have a weak inverse association with breast cancer risk. However, recent data that includes molecular subtyping information provides evidence of a possible stronger inverse association for hormone-negative breast tumors.(6-9,20,21) In the AMBER study, the inverse association of breastfeeding was most pronounced for younger-onset ER-negative and triple-negative breast tumors. In fact, for such tumors, analyses demonstrated that extended breastfeeding could reduce the adverse risks associated with parity, which has also been seen in other studies that included African-American women.(9,11) Our results revealed similar associations given that extended breastfeeding appeared to largely counteract the adverse relationship with multiparity among younger women with ER-negative tumors.

Recent studies assessing associations by molecular subtypes using IHC and mRNA expression profiling have shown increased risk with parity that may predominate for triple-negative or basal-like breast tumors.(20,24) In our study, the modifications in risk associations between parity and breastfeeding by age reflected different temporal trend patterns by birth cohorts in cases and controls: the rate of decrease in number of births was faster for controls than cases in early birth cohorts (i.e., older women); a decreasing trend of breastfeeding months per pregnancy in early birth cohorts was seen in controls but not in cases. Given that multiparity and increased breastfeeding are inversely associated with later-onset breast cancers (with somewhat stronger associations with ER-positive tumors), if the observed temporal trends of decreasing parity and breastfeeding continue, they are likely to result in an increased incidence of later-onset breast cancer.(13) This indicates the importance of public health measures to maintain high rates of breastfeeding,(25) which could potentially attenuate the projected increase in risk due to changes in reproductive patterns and demographics.(13,26)

Older age at first birth has been associated with increases in breast cancer risk in numerous studies, particularly for ER-positive tumors.(8,10,20,27,28) The AMBER consortium also found increased risks for older ages at first birth for ER-positive but not for ER-negative tumors. Our data were consistent with these findings, suggesting that this association may be stronger or limited to early-onset ER-positive breast cancer cases.(6) However, in African populations, this is a difficult exposure to assess given that few women actually delay their first births until truly late ages. With increasing adoption of westernized lifestyles and access to birth control, continued monitoring of maternity data are needed to determine if ages at first birth continue to increase.

In spite of the observed trends in reproductive patterns toward westernization, our study population still maintained higher parity and breastfeeding frequencies compared with other populations. The reproductive patterns in our study are consistent with recent nationally representative surveys.(29,30) For example, the decline in fertility rate from 6.4 in 1988 to 3.9 in 2017 reported in surveys by the Ghana Maternal Health Survey ages 15–49 years is similar to the decline in average number of live births in our control population from 6.4 to 3.3 for women born in 1945 (i.e. 43 years old in 1988) and 1975 (i.e. 42 years old in 2017).(29) Median breastfeeding months per pregnancy were 17 to 18 months in our study controls and in a 2011 survey median months breastfeeding were 17.4 and 17.9 months for Greater Accra and Ashanti regions, respectively.(30) The strong inverse associations of these factors with late onset, mostly ER-positive tumors, together with a lack of population-based screening, are likely important factors contributing to historically low incidence of late onset ER-positive breast cancers. In contrast, for early-onset cancers, higher parity was directly associated with ER-negative disease in our study. It is doubtful, however, that high parity explains the higher incidence of ER-negative early-onset cancers in our population given the high prevalence of breastfeeding, which appeared to offset the higher risk from multiparity. Instead, the younger demographics in Ghana and other sub-Saharan African countries probably explains the higher proportion of these early-onset cancers compared with populations of European ancestry.(3) It may be that rather than a population with an “excess” of early-onset ER-negative cancers that there could be fewer diagnoses of late onset ER-positive breast cancer compared with other populations, as suggested in other studies.(31) To specifically address this, further studies comparing age-incidence rates of breast cancer subtypes in Africa are needed, similar to U.S. studies that have addressed racial differences by age.(32)

Age at menarche has been inversely associated with risk in European ancestry populations.(33) In the studies of African-American women, later ages at menarche were inversely associated with breast cancer regardless of hormone receptor status.(5,9) In contrast, we observed no such relationship. The median age of menarche of 15 years in Ghanaian women is quite different from the reported age of 12 years among African-American women, with our study having limited variation in ages at menarche. Increased nutrition has been suggested to lower the age at menarche; this variable could reflect early exposures that may differ between populations (e.g. early adolescent weight).(34) In addition, a substantial number of women in our study could not recall their ages at menarche, suggesting that measurement error could have impacted our ability to assess relationships reliably.

Strengths of this study are the population-based design, detailed risk factor assessment, and tissue collection for quality assessment of IHC markers to examine etiologic heterogeneity in sub-Saharan Africa. A limitation is that although IHC data can be used as a proxy for molecular subtypes, mRNA expression assays are required to classify previously described intrinsic molecular subtypes, especially HER2-enriched and luminal B subtypes. Further, although our study is one of the largest breast cancer epidemiological studies conducted in sub-Saharan Africa, analyses by age and subtypes resulted in small numbers within strata of these critical factors.

Our study indicates that while reproductive factors showed important temporal trends and distinct distributions compared with African-American or European ancestry populations, their associations with breast cancer risk were generally consistent with those observed in these populations. Our data support the importance of breastfeeding to prevent early-onset ER-negative breast cancer associated with multiparty and the longer-term protection of parity and breastfeeding for later-onset breast tumors, irrespective of their ER status. Further studies including more detailed molecular characterization of tumors and additional risk factors may provide additional insights into breast cancer etiology in sub-Saharan Africa.

## Data Availability

As these data are from patients, summary statistics are available but individual level data require IRB/human subjects approvals and data transfer agreements.

## List of abbreviations

ER: estrogen receptor
OR: odds ratios
CI: confidence interval
PR: progesterone receptor
HER2: human epidermal growth factor receptor 2
AMBER: African American Breast Cancer Epidemiology and Risk
IHC: immunohistochemical
NCI: National Cancer Institute
ABCS: African Breast Cancer Study

## Funding

This work was supported by Intramural program of the National Cancer Institute.

## Acknowledgments

We are grateful to all women who agreed to participate in the study and provided information and biospecimens.

## Author contributions

Conception & Design of the study: JDF, BDL, LE, NT, EA, JNCL, JY, BW, BA, MD, SW, KN, FA, DA, SH, TA, MGC, LAB; Data collection: JDF, BDL, LE, NT, EA, JNCL, JY, BW, BA, MD, SW, KN, FA, DA, SH, TA, MGC, LAB; Interpretation of data JDF, BDL, LE, NT, EA, JNCL, JY, BW, BA, MD, SW, KN, FA, DA, SH, TA, MGC, LAB; Drafting of the manuscript: JDF, BDL, LAB, MGC; Revised work and provided important intellectual content: JDF, BDL, LE, NT, EA, JNCL, JY, BW, BA, MD, SW, KN, FA, DA, SH, TA, MGC, LAB.

## Conflict of Interest

none declared

## Ghana Breast Health Study team

Ghana Statistical Service, Accra, Ghana: Dr Robertson Adjei and Dr Lucy Afriyie. Korle Bu Teaching Hospital, Accra, Ghana: Dr Anthony Adjei, Dr Florence Dedey, Dr Verna Vanderpuye, Victoria Okyne, Naomi Ohene Oti, Evelyn Tay, Dr Adu-Aryee, Angela Kenu and Obed Ekpedzor. Komfo Anoyke Teaching Hospital, Kumasi, Ghana: Marion Alcpaloo, Isaac Boakye, Bernard Arhin, Emmanuel Assimah, Samuel Ka-chungu, Dr Joseph Oppong and Dr Ernest Osei-Bonsu. Peace and Love Hospital, Kumasi, Ghana: Prof Margaret Frempong, Emma Brew Abaidoo, Bridget Nortey Mensah, Samuel Amanama, Prince Agyapong, Debora Boateng, Ansong Thomas Agyei, Richard Opoku and Kofi Owusu Gyimah. Memorial Sloan Kettering Cancer Center, NY, USA: Dr Lisa Newman. National Cancer Institute, Bethesda, MD, USA: Maya Palakal and Jake Thistle. Westat, Inc.: Michelle Brotzman, Shelley Niwa, Usha Singh and Ann Truelove. University of Ghana: Prof Richard Biritwum.

## Notes

### Competing Interest Statement

The authors have declared no competing interest.

### Clinical Protocols

https://onlinelibrary.wiley.com/doi/full/10.1002/ijc.30688

### Author Declarations

All relevant ethical guidelines have been followed and any necessary IRB and/or ethics committee approvals have been obtained.

Any clinical trials involved have been registered with an ICMJE-approved registry such as ClinicalTrials.gov and the trial ID is included in the manuscript.

